# Long-term risk of cardiovascular disease after assisted reproductive technology and infertility

**DOI:** 10.64898/2026.05.18.26353477

**Authors:** Angelo G. Mezzoiuso, Peter Henriksson, Márta Radó, Kenny Rodriguez-Wallberg, A. Sara Öberg

## Abstract

**Background:** The use of Assisted Reproductive Technology (ART) is increasing worldwide. These treatments involve ovarian stimulation to enable multiple follicle recruitment, hence inducing supraphysiological estrogen levels. While most long-term follow-up of women undergoing ART has concerned cancer incidence, the long-term safety regarding cardiovascular and metabolic diseases remains under-explored. This study was performed to assess the risk of acute myocardial infarction, cerebral ischemic conditions, intracranial hemorrhage, type 2 diabetes mellitus, heart failure, aortic aneurysm or dissection, and chronic kidney disease in women that conceived with ART, and to investigate the role of the underlying infertility and its risk factors.

**Methods and Findings:** Swedish national registers allowed us to follow a nationwide cohort of 380,756 women from their first birth between 1992 and 2002 until the end of 2023. The safety of ART was evaluated by comparing women with infertility who conceived with and without ART, while adjusting for baseline differences in age, body mass index, country of origin, socioeconomic factors, pre-existing comorbidity, smoking and year. The role of infertility was additionally explored by comparing all women with and without infertility adjusting for age, as well as the aforementioned baseline characteristics. Cumulative risks were plotted using inverse-probability weighted Kaplan-Meier curves. To facilitate the comparison of groups we also estimated risk differences and ratios at 10-, 20-, and 30-years of follow-up. Use of ART was not associated with cardiovascular disease except for an excess risk of cerebral ischemic conditions, with a 30 year risk ratio of 1.43 (1.09; 1.89). With the exception of cerebral ischemic conditions, intracranial hemorrhage, aortic dissection, and chronic kidney disease, women with a history of infertility exhibited consistently higher risk of all outcomes, adjustment for differences in baseline characteristics explained some but not all of these elevated risks.

**Conclusions:** With the exception of ischemic cerebral conditions, the findings provide reassurance regarding the long-term cardiometabolic safety of ART use, while adding to the growing literature suggesting that infertility can act as a marker of women’s cardiovascular and metabolic disease.

**AUTHOR SUMMARY:** *Why Was This Study Done?:*

- Assisted reproductive technology is an increasingly common infertility treatment, accounting for up to 5% of children born per year in some countries.
- The treatment often exposes women to high doses of hormones, which raises concerns about potential negative effects on the cardiovascular system.
- It is currently unclear whether use of assisted reproductive technology increases a woman’s long-term risk of cardiovascular diseases.

*What Did the Researchers Do and Find?:*

- We conducted a study including all women who gave birth in Sweden from 1992 to 2002 [380,756], with ability to track the occurrence of heart disease and stroke through 2023.
- We compared women who conceived with and without assisted reproductive technology after experiencing infertility, with adjustment for differences in background characteristics. We also compared overall risks to women with and without infertility.
- Use of assisted reproductive technology was associated with a 43% higher risk of cerebral ischemic conditions, but not with any other studied outcomes. After 30 years follow-up, women with infertility were found at 36% higher risk of acute myocardial infarction, 34% higher risk of type 2 diabetes, and 25% higher risk of heart failure, compared to women with similar background characteristics.

*What Do These Findings Mean?:*

- While the fidings are largely reassuring with respect to long-term cardiovascular health following use of ART, an excess risk of cerebral ischemic conditions warrants further consideration.
- The results suggest that women with infertility are at elevated risk of cardiovascular issues later in life.
- Limitations of the study include the relatively small number of women treated with these technologies and a lack of data on women who underwent treatment which did not result in birth.

## INTRODUCTION

Assisted reproductive technology (ART) is an increasingly utilized and effective treatment for infertility [1], with recent global data showing over 3.3 million ART cycles being performed across 83 countries in 2018 [2]. In ART, women undergo hormonal stimulation to develop several follicles from which oocytes can be retrieved for in-vitro fertilization (IVF), sometimes using intracytoplasmic sperm injection (ICSI). After a few days of culture, embryos are transferred to the uterus and/or frozen for later transfer. The full extent to which ART treatment protocols, and the controlled hyperstimulation of the ovaries, may influence women’s cardiovascular health is not clear, especially not in the long-term [3].

Sex hormones play a complex and still not well understood role in cardiovascular health. It has for example been suggested that declining estrogen after menopause [4], and increased androgens in conditions like polycystic ovarian syndrome could be linked to higher cardiovascular risk in women [5]. In the last years, growing evidence suggests that women who undergo ART are at increased risk of cardiovascular morbidity in the short term, particularly for venous thromboembolism, which has also been linked to the hyperstimulation of the ovaries [6–8]. Furthermore, a cohort study with an average follow-up of 9 years observed an elevated risk of hypertension and possibly also of stroke in women that gave birth following ART [9]. Seeing how hypertension contributes to cardiovascular disease through chronic exposure, extended follow-up will be necessary to observe its full consequences. Because pregnant women are a population already at risk in the short-term, it is important to understand if a treatment that further increases this risk could also affect their long-term risk of cardiovascular diseases.

Until now, the long-term cardiovascular safety of ART has received limited attention in research. A recent meta-analysis on the topic included only six studies and, due to the large heterogeneity, was unable to draw conclusions beyond the necessity of more well-characterized studies [10]. With the exception of a recent study using Nordic register data, previous efforts have also been characterized by relatively short follow-up. As cardiovascular disease risk factors may act for long periods before disease onset, too short follow-up could underestimate the real impact and sometimes even introduce bias [11]. Another largely overlooked challenge is to disentangle the role of ART from that of its indication, since factors related to infertility, such as specific diseases, could be responsible for cardiovascular disease development. This distinction is crucial for accurately evaluating the safety of ART as well as understanding the overall risk to women with infertility [12–14].

This nationwide cohort study, based on prospectively collected register data, examined the long-term risk of specific cardiovascular diseases in women that experienced infertility and potential use of ART prior to their first birth, with comprehensive follow-up across 30 years allowing evaluation of long-term cardiovascular outcomes beyond the timeframes of most previous studies. Multiple comparisons and adjustment for measured background characteristics were used to inform on the roles of ART and infertility.

## METHODS

### Study hypotheses

This study was designed to answer the following questions: is use of ART associated with elevated risk of cardiovascular disease overall (primary descriptive aim); could observed risks be explained by underlying characteristics related to infertility rather than exposure to the treatment (primary causal aim); and finally to what extent are women with primary infertility at elevated risk of cardiovascular disease compared to those without (secondary descriptive aim).

### Data sources

This cohort study was conducted using data from multiple Swedish population-based registers, which were linked at the individual level using the unique personal identity number assigned to all residents [15]. The study is based on the Medical Birth Register (MBR), which was established in 1973 and collects antenatal and delivery data on all hospital-based births [16]. By 10-12 weeks of pregnancy, most women are enrolled in antenatal care provided by midwives [17]. Since 1982 the standardized interview has included questions about involuntary childlessness, time to pregnancy, and since 1995 also potential use of fertility treatment (ART, intrauterine insemination, surgery, or ovulation induction). Information from subsequent gestational visits is recorded until the delivery, which almost always occurs in a hospital. Public and private health centers involved in antenatal care and delivery are legally required to send select information to the National Board of Health and Welfare. The datasets are merged, undergo quality checks, and are then released to the MBR. The register consistently covers 97-99% of all births in Sweden [16]. Since the first birth from ART in Sweden in 1982, the MBR started collecting information from all IVF clinics in Sweden, including the type of fertilization method (IVF/ICSI) and the use of fresh or frozen embryos.

The National Patient Register includes all completed inpatient admissions with nationwide coverage since 1987. It also contains data on patients treated in specialized outpatient care since 2001. A primary and up to seven contributing diagnoses are coded using the Swedish version of the International Classifications of Diseases (ICD) in use at the time of the visit [18]. Procedures are recorded using the prevailing Swedish code system. The ICD is also used to record the main and up to eight contributing causes of death in the Cause of Death Register since 1952 [19]. The quality of information depends on the quality of the ascertainment from the physician who compiled the death certificate. For cardiovascular disease, the agreement between the official cause of death and the re-evaluated certificates has been found at 87-88% for chapter level and at 55% for greater detail [20]. Finally, registers kept by Statistics Sweden provide core demographic information such as country of origin, dates of migration to and from Sweden, attained level of education, and socioeconomic index.

### Study Population

All 874,869 women who had their first birth recorded in the MBR between 1982 and 2002 were eligible for the study. The selection of the study population is illustrated in Figure 1. To allow all women the same eligibility to enrol in antenatal care at the start of pregnancy, 17,772 women who migrated to Sweden within 280 days from the delivery were excluded. Because some identity numbers of deceased individuals have been reassigned to new Swedes, another 85,835 women were excluded to avoid incorrect attribution of cardiovascular events to individuals that did not experience them. To further ensure that only incident cases were considered, 4,979 women who had a recorded diagnosis of any of the outcomes of interest prior to the start of follow-up were excluded. Finally, to allow equal opportunity of ART and focus on primary infertility we elected to align time zero with the first birth, and consequentially excluded 2,319 women who had undergone ART for a birth that was not their first, to avoid exposure misclassification.

**Figure 1.**
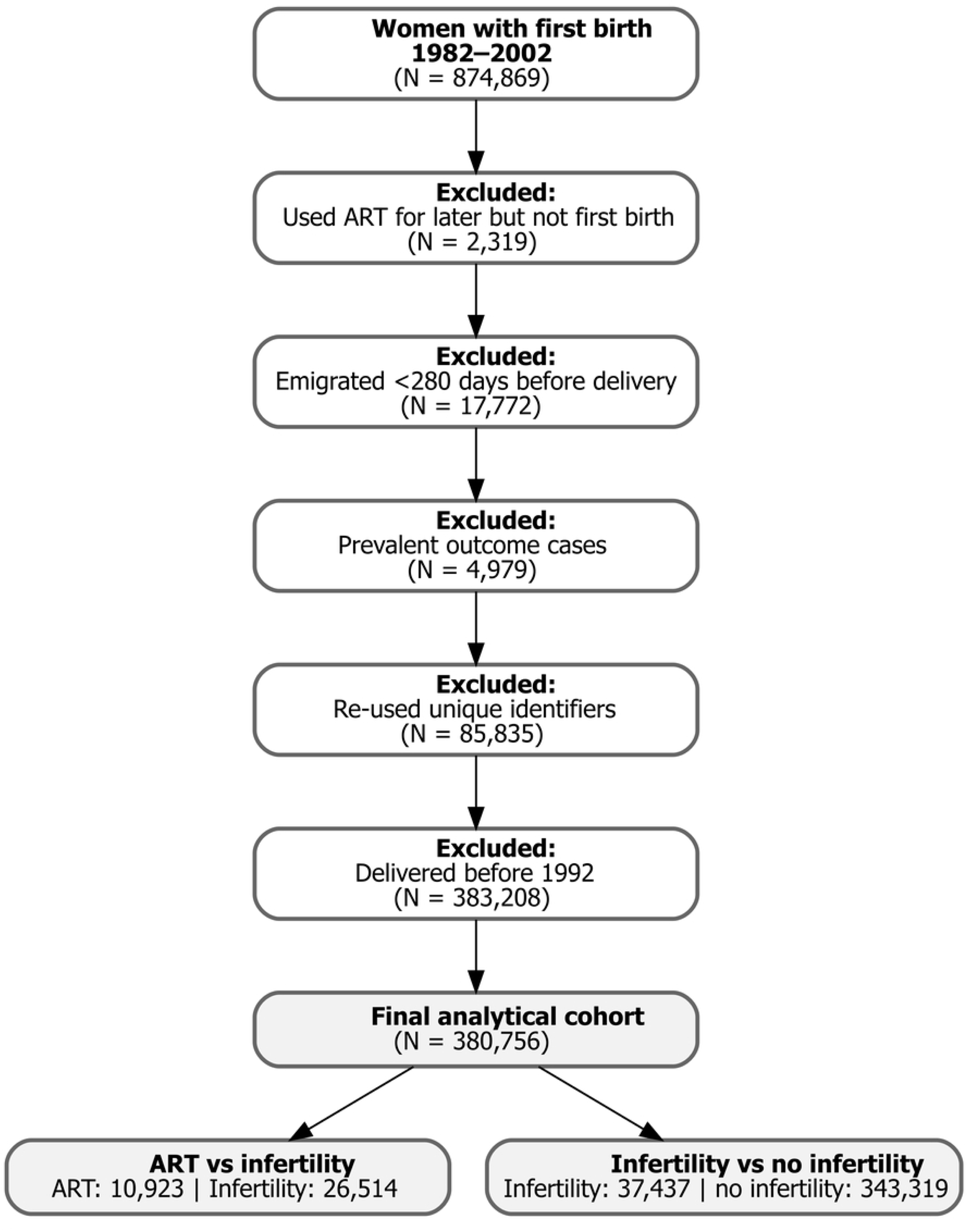
Study flowchart and exclusions.

Finding very few, and likely highly selected, women exposed to ART in the first years of its implementation, it was ultimately deemed more appropriate to restrict the study period to births from 1992. The remaining 380,756 women could be followed from their estimated conception date until whichever came first of the cardiovascular outcome of interest (i.e., regardless of whether any of the other outcomes had occurred previously), emigration from Sweden, death, or the end of follow-up on December 31, 2023.

### Exposure and outcomes

Women’s use of ART was primarily identified using IVF clinic reports to the MBR, complemented with women’s self-report under the assumption it could capture potential treatments made abroad. Infertility, defined as failure to achieve pregnancy after more than 12 months of regular unprotected sex, was identified from women’s self-reported time-to-pregnancy at the first antenatal visit. Diagnostic and procedure codes for infertility and fertility treatment recorded in the patient register were also considered but made limited contributions considering outpatient care was only available in the very end of the study period (for a detailed list of codes see S1 Table). To cover a spectrum of cardiovascular diseases, the following outcomes were considered: acute myocardial infarction, cerebral ischemic events (including cerebral infarction, occlusion and stenosis of precerebral and cerebral arteries), intracranial hemorrhage, type 2 diabetes mellitus, heart failure, aortic aneurysm or dissection, and chronic kidney disease. For each respective outcome, the first recorded diagnosis in inpatient or outpatient specialist care was the primary source of identification, complemented by a fatal event recorded as a cause of death when applicable. Given the heterogeneity of heart failure, aortic aneurysm or dissection, and chronic kidney disease, we applied selection criteria to capture only those cases most plausibly linked to cardiovascular disease rather than to other underlying causes. The specific ICD codes and selection criteria used across ICD editions 8 (before 1987), 9 (1988–1996), and 10 (from 1997) are detailed in S2 Table.

### Covariates

A range of measured background characteristics were considered for their putative role as risk factors for infertility and ART use as well as cardiovascular disease development, and their distribution in relation to infertility and ART status are summarized in the descriptive Table 1. Relevant confounders for the causal evaluation of ART safety were identified based on their subject-matter informed hypothesized causal relationships with infertility, fertility treatments, and cardiovascular diseases (S1 Fig). Ultimately we selected to account for baseline differences in age, body mass index (BMI), and calendar year (treated as continuous variables), and country of origin, education, socioeconomic index, smoking status, and self-reported chronic conditions (treated as categorical variables).

**Table 1.** Characteristics of women prior to their first childbirth in Sweden between 1992 and 2002.

|  | No ART |  | ART |
| --- | --- | --- | --- |
|  | No infertility | Infertility | ART |
|  | N= 343,319 | N= 26,514 | N= 10,923 |
| **Characteristics** |  |  |  |
| Age in years, Mean (SD) | 26.0 (4.6) | 28.9 (4.6) | 32 (4.0) |
| Year of birth |  |  |  |
| 1992 - 1995 | 139,405 (40.6%) | 10,711 (40.4%) | 2,594 (23.7%) |
| 1996 - 1998 | 84,148 (24.5%) | 6,726 (25.4%) | 3,207 (29.4%) |
| 1999 - 2002 | 119,766 (34.9%) | 9,077 (34.2%) | 5,122 (46.9%) |
| BMI (kg/m<sup>2</sup>), Mean (SD) | 23.6 (3.8) | 24.3 (4.5) | 24.2 (3.9) |
| BMI categories, n (%) |  |  |  |
| Underweight | 9,116 (2.7%) | 607 (2.3%) | 144 (1.3%) |
| Normal range | 203,008 (59.1%) | 15,035 (56.7%) | 6,003 (55.0%) |
| Overweight | 57,403 (16.7%) | 5,190 (19.6%) | 2,272 (20.8%) |
| Obese | 19,156 (5.6%) | 2,501 (9.4%) | 773 (7.1%) |
| *Missing* | *54,636 (15.9%)* | *3,181 (12.0%)* | *1,731 (15.8%)* |
| Socioeconomic Index, n (%) |  |  |  |
| Low | 64,312 (18.7%) | 5,892 (22.2%) | 2,136 (19.6%) |
| Middle-low | 105,407 (30.7%) | 9,482 (35.8%) | 4,192 (38.4%) |
| Middle-high | 31,801 (9.3%) | 3,534 (13.3%) | 1,924 (17.6%) |
| High | 13,694 (4.0%) | 1,593 (6.0%) | 961 (8.8%) |
| *Missing* | *128,105 (37.3%)* | *6,013 (22.7%)* | *1,710 (15.6%)* |
| Country of origin |  |  |  |
| Sweden | 330,434 (96.2%) | 25,691 (96.9%) | 10,562 (96.7%) |
| Other European countries | 6,745 (2.0%) | 523 (2.0%) | 253 (2.3%) |
| Non-European countries | 6,140 (1.8%) | 300 (1.1%) | 108 (1.0%) |
| Highest attained education, n (%) |  |  |  |
| Lower Secondary | 47,452 (13.8%) | 2,505 (9.5%) | 741 (6.8%) |
| Upper Secondary | 187,115 (54.5%) | 15,017 (56.6%) | 5,538 (50.7%) |
| Post-secondary | 102,514 (29.9%) | 8,911 (33.6%) | 4,618 (42.3%) |
| *Missing* | *6,238 (1.8%)* | *81 (0.3%)* | *26 (0.2%)* |
| Smoking in pregnancy, n (%) |  |  |  |
| Non-smoker | 274,850 (80.1%) | 22,224 (83.8%) | 9,392 (86.0%) |
| Smoker | 50,460 (14.7%) | 3,748 (14.2%) | 876 (8.0%) |
| *Missing* | *18,009 (5.2%)* | *542 (2.0%)* | *655 (6.0%)* |
| Previous/chronic diseases, n (%) |  |  |  |
| Asthma | 18,959 (5.5%) | 1,464 (5.5%) | 451 (4.1%) |
| Diabetes | 240 (0.1%) | 29 (0.1%) | 12 (0.1%) |
| Hypertension | 773 (0.2%) | 83 (0.3%) | 23 (0.2%) |
| Epilepsy | 1,135 (0.3%) | 93 (0.3%) | 30 (0.3%) |
| Chronic kidney diseases | 1,389 (0.4%) | 144 (0.5%) | 40 (0.4%) |
| Systemic lupus erythematosus | 296 (0.1%) | 18 (0.1%) | <10 (0.1%) |
| Inflammatory bowel disease | 1,436 (0.4%) | 147 (0.5%) | 78 (0.7%) |
| Follow-up, median (Q1-Q3) | 29.3 (24.5 – 30) | 28.2 (24.5 – 30) | 25.9 (23.6 – 28.8) |

Footnote: Country of origin, education and socioeconomic index come from demographic records at Statistics Sweden, and all other variables from the Medical Birth Register (BMI, smoking and chronic conditions are based on self-reported responses to the standardized interview at the first antenatal visit)

### Statistical analysis

Potential differences in background characteristics between the mutually exclusive groups of all women that conceived with ART, after infertility, and no infertility respectively, were examined with Student’s t-tests, Mann-Whitney tests, and Chi-square tests.

The primary analysis concerned the safety of ART. This causal aim was achieved by comparing women who conceived with and without ART after having experienced infertility before their first birth, while adjusting for a priori identified confounders to account for potential systematic differences in baseline characteristics. A secondary analysis was performed to additionally inform on the role of primary infertility. This descriptive aim compared all women with a record of infertility prior to their first birth to all other women, adjusting only for age to capture the overall risk associated with infertility, and then also for all previously noted baseline characteristics to assess their influence.

Kaplan-Meier failure curves were estimated and plotted for each outcome to allow comparison of the full risk profiles over time. Groups were additionally compared by estimating the 10-, 20-, and 30-year cumulative risk difference and risk ratio with corresponding 95% confidence intervals. Inverse probability of treatment weights (IPTW) were used to account for relevant differences between contrasted groups. Logistic regression models were used to obtain propensity scores (probability of treatment given covariates) which were then used to assign each woman a weight corresponding to the inverse probability of being treated (if treated) or not treated (if not treated). In the propensity score models used to create the weigths, continuous variables (age, BMI, and calendar year) were modeled using restricted cubic splines with 3 degrees of freedom to allow for non-linear associations with the treatment assignment. Weights were stabilized by the marginal probability of exposure, all winsorized to explore the role of extreme weights, and truncation at the 1^st^ and 99^th^ percentile reduced the maximum IPTW from 18.37 to 2.57 (S3 Fig.) Mirror density plots were employed to assess the degree of overlap between treatment groups and standardized mean differences were used to determine the balance of baseline characteristics between the treated and not treated before and after weighting, with SMD thresholds of <0.1 and <0.2 considered optimal and acceptable indications of balance. These maintained acceptable balance with all SMDs <0.2 (S4 Table) and propensity score distributions showed near-perfect overlap between groups (S4 Figure).

Multivariable imputation by chained equations using the fully conditional specification [21] was employed to impute information when such was missing for covariates in the fully adjusted IPTW model (BMI, education, socioeconomic index, smoking and country of origin) in 20 data sets, and the estimates from these were pooled using Rubin’s rules [22]. Diagnostic assessments of the procedure confirmed excellent model performance, with Markov Chain Monte Carlo trace plots demonstrating convergence (S2 Fig, Panel A) and distributional comparisons showing close aligment between observed and imputed data (S2 Fig, Panel B-C; S3 Table).

### Sensitivity analysis

First, to allow assessment of the influence of weight truncation, the fully adjusted risk estimates for the main outcomes were obtained using also the non-winsorized IPTW. Second, to address potential residual confounding influence of severity (in infertility and/or its risk factors) in the primary analysis of ART safety, a sensitivity analysis assessed the women who conceived through ICSI separately from standard IVF. Because ICSI was strictly reserved for male factor infertility in this early period in Sweden, the subset of women in which this fertilization technique was used could be expected to have fewer infertility-related risk factors than those undergoing standard IVF. Under the null hypothesis (no causal effect of ART itself), the women who conceived with ICSI would therefore be expected at similar or even lower risk than women with infertility who conceived without ART. Due to the later implementation of ICSI, the analysis was restricted to women whose first birth occurred in 1994 or later. This study is reported as per the Strengthening the Reporting of Observational Studies in Epidemiology (STROBE) guideline (S1 Checklist) [23]. The performed analyses align with what was originally planned when the study was designed, with the following notable exceptions: the secondary analysis to quantify overall risk to women with primary infertility was amended to further strengthen the contribution of the study. Additionally, the earliest years of ART implementation were eventually excluded based on the number of women exposed being so few and possibly highly selected. All analyses were performed using the software SAS 9.4 version and R version 4.3.1.

### Ethics Statement

This study was approved by the Swedish Ethical Review Authority (Etikprövningsmyndigheten; DNR 2013/1849-31/2, DNR 2018/386-32, DNR 2019-01529). The study used de-identified registry data, and the Authority waived informed consent in accordance with Swedish law and GDPR for research using pseudonymized data.

### Data availability

Individual-level data from the register-linkage used in the study cannot be made publicly available under current legislation for data protection (General Data Protection Regulation, Swedish law (SFS 2018:218), the Swedish Data Protection Act, the Swedish Ethical Review Act and the Public Access to Information and Secrecy Act). Researchers who wish to replicate the study can request approval from the Swedish Ethical Review Authority to apply for data extraction directly from the respective register holders; the Swedish National Board of Health and Welfare, Statistics Sweden. The data dictionary and analytic code are available in a repository at https://github.com/Epidemiologit/Long-term-risk-of-cardiovascular-disease-after-assisted-reproductive-technology-and-infertility-Code. This repository will be archived in Zenodo to provide a permanent DOI upon article publication.

## RESULTS

### Background characteristics

The study population included 380,756 women, of whom (7.0%) had experienced infertility but ultimately conceived without ART, and 10,923 (2.9%) conceived with ART. The background characteristics of these three mutually exclusive groups, defined by infertility and use of ART, are presented in Table 1. At the time of the first birth, women who conceived with ART were on average older (mean age 32 years) compared to women with infertility who conceived without ART, (29 years) and all other women with no known infertility (26 years). Additionally, the women who had used ART were more likely to have conceived toward the end of the study period, less likely to be smokers and more likely to have post-secondary education compared to non-treated women with and without infertility (who were generally more similar with respect to these characteristics). No substantial differences were found regarding the prevalence of the chronic conditions surveyed at the first antenatal visit, except for a lower prevalence of asthma and higher prevalence of inflammatory bowel disease among treated women (asthma 4.1% compared to 5.5% in all other women, IBD 0.7% compared to 0.5% and 0.4% in women with and without infertility). Because use of ART was much more common toward the end of the study period, women that gave birth after ART had a slightly shorter median follow-up (25.9 years) compared to 28.2 and 29.3 years in women with and without infertility respectively). In all ensuing estimation of risk, we censored the follow-up at 30 years post conception (and 25 years in the analysis of ICSI), to allow reasonable numbers of women still at-risk.

The main findings for the primary (ART) and secondary (infertility) analysis are presented in Figures 2 and 3 showing the Kaplan-Meier estimated failure curves for acute myocardial infarction, type 2 diabetes, ischemic cerebral conditions, heart failure and intracranial hemorrhage, and tables showing the corresponding risk comparisons at 10-, 20-, and 30-years. Additional findings for chronic kidney disease and aortic aneurysm or dissection are provided in separate figures.

**Figure 2.**
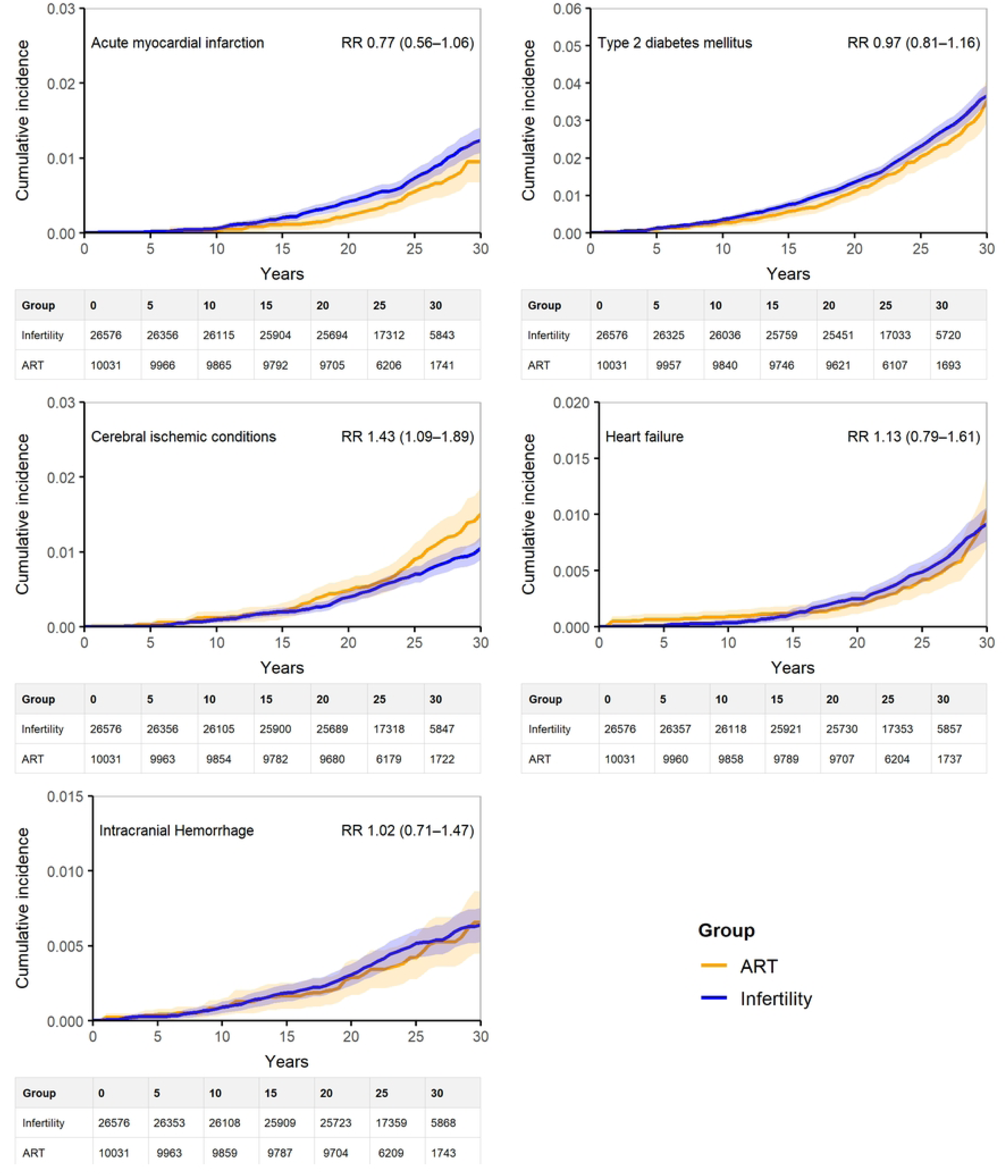
Kaplan–Meier failure curves of the main cardiovascular outcomes for the fully-weighted comparison of ART and no ART among women with primary infertility.

**Figure 3.**
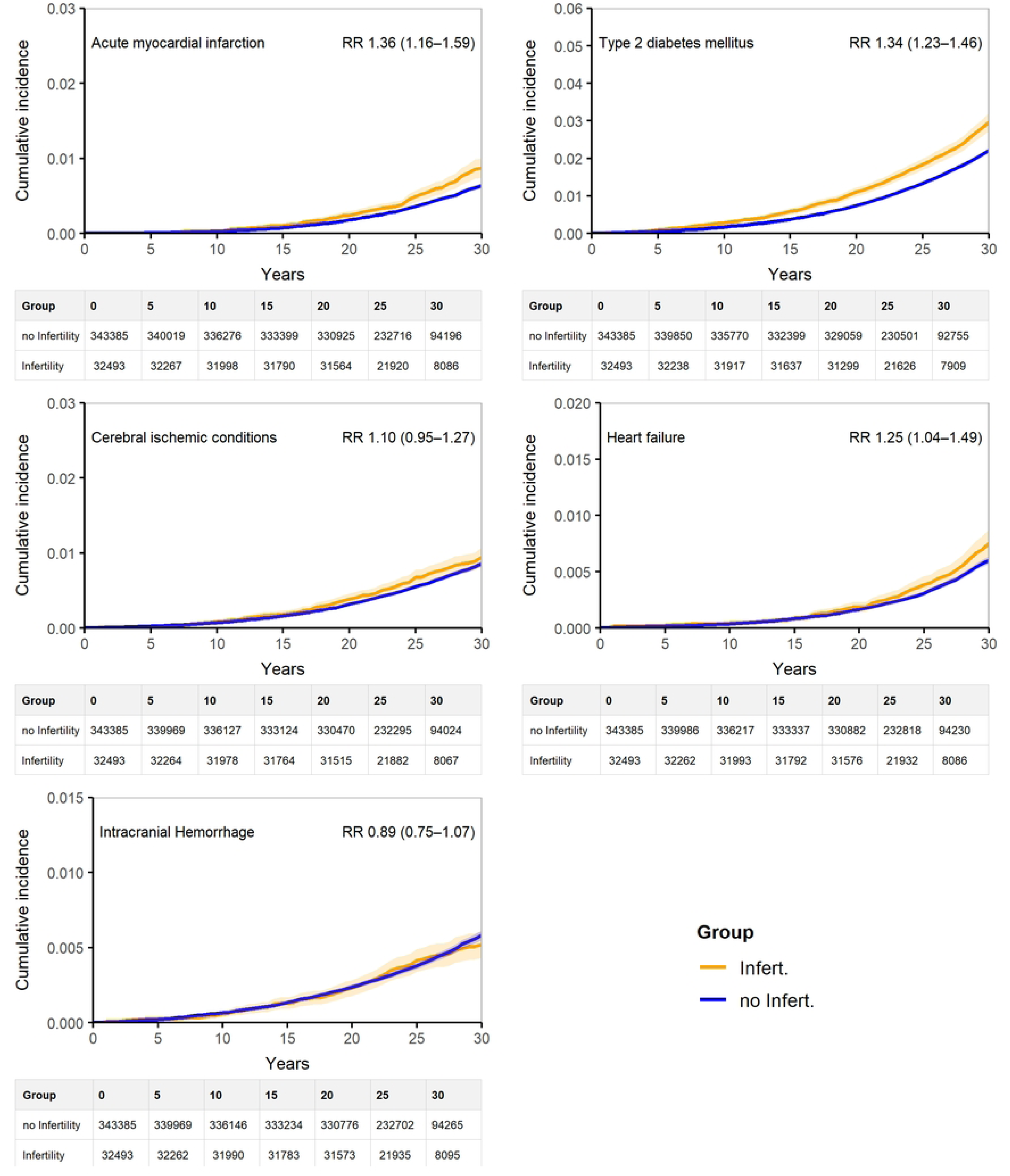
Kaplan–Meier failure curves of the main cardiovascular outcomes for the fully-weighted comparison of women with and without primary infertility.

### Primary analysis

In the causal comparison of women that conceived with and without ART after experiencing primary infertility while adjusting for background characteristics, the women who conceived with ART were not at excess risk for any of the outcomes except for gradually developing elevated risk of cerebral ischemic conditions toward the end of the follow-up with a risk difference of 0.46% (0.07%; 0.84%) and a relative risk of 1.43 (1.09; 1.89) (Figure 2, S5 Table).

### Secondary analysis

The descriptive comparison of all women who experienced infertility prior to their first birth to similar-aged women in the cohort revealed consistently elevated risk of all of the main outcomes except for cerebral ischemic conditions, intracranial hemorrhage, aortic aneurysm/ dissection or the selected chronic kidney disease subtypes, chosen based on their known association with cardiovascular risk factors (S6 Fig).

Further adjustment for differences in baseline characteristics appeared to explain some, but not all of the observed excess risk (Figure 2). For acute myocardial infarction, this corresponded to an attenuation of the 30 year risk difference from 0.29% (0.16; 0.43) to 0.23% (0.10%; 0.36%), and of the relative risk from 1.47 (1.26; 1.71) to 1.36 (1.16; 1.59)(S6-S7 Tables). Similarly, the culminating risk difference of 1.24% for type II diabetes (0.97%; 1.52%) was attenuated to 0.75% (0.50%; 1.00%) and the corresponding risk ratio from 1.58 (1.45; 1.71) to 1.34 (1.23; 1.46). The estimated excess risk of heart failure after 30 years went from 0.21% (0.07%; 0.34%) to 0.15% (0.02%; 0.28%) risk difference and from 1.35 (1.14; 1.61) to 1.25 (1.04; 1.49) relative risk. While slightly elevated risks of cerebral ischemic conditions were observed across follow-up, these could not consistently be distinguished from the null, especially not after adjustment (30-year risk ratio 1.15 (1.00; 1.13) and 1.10 (0.95; 1.27) respectively (S6-7 Tables).

### Sensitivity analyss

Side-by-side comparison of the fully adjusted risk estimates based on winsorized and non-winsorized IPTW showed materially overlapping results (S5 Table). Also the results of only considering women who conceived with ICSI technique were consistent with the overall findings. With the exception of cerebral ischemic conditions, this group appeared at similar or lower risk than women with infertility who conceived without ART (S8 Fig). Notably, while the risk of cerebral ischemic conditions appeared slightly elevated it could not be distinguished from that of the comparison group.

## DISCUSSION

### Summary of results

In this nation-wide cohort study following women for 30 years from their first birth, the women who had used ART were not at increased risk of any of the studied cardiovascular diseases except cerebral ischemic conditions, compared to all other women with infertility. We additionally observed that over time and regardless of potential ART use, the women who experienced infertility prior to the first birth developed an excess risk of acute myocardial infarction, type II diabetes, and heart failure, in comparison to all other women. While these findings are largely reassuring regarding the long-term cardiovascular safety of ART, they add to the growing literature noting an excess risk of cerebral ischemic conditions and also highlighting infertility as a consistent cardiovascular risk factor in women.

### Comparison with previous studies

Our results are consistent with other large population-based studies, which have mostly not found women giving birth after ART at elevated long-term risk of major cardiovascular events.[3,24] Shorter spanning studies in Canada, Sweden and across the Nordic countries (median follow-up to 11 years) have generally reported no excess risk for major outcomes, with some evidence of more hypertension and possibly also an elevated risk of stroke. In the largest study to date, including 2.5 million women from four Nordic countries, ART was not associated with cardiovascular outcomes except a slight excess risk of stroke in women giving birth after frozen embryo transfer [24]. Our study adds to these largely reassuring findings paired with a risk elevation of cerebral ischemic conditions. Finally, while a recent meta-analysis of 10 studies covering over 500,000 exposed women, has also provided reassurance with respect to long-term risks of coronary heart disease, stroke, venous thromboembolism, hypertension diabetes and heart failure, we note that they considered also non-ART interventions such as intrauterine inseminations and ovulation induction as exposure to ART [25].

This study substantially extends the available follow-up of most previous studies, allowing a substantial proportion of women to reach an age when cardiovascular outcomes are no longer rare or atypical. By considering events from conception onward, we furthermore describe the full risk profile from the time of exposure. This particularly contrasts with the largest study to date, which started follow-up 2 years postpartum, effectively omitting both pregnancy-related and early postnatal cardiovascular risk. Our outcome consideration further allowed cerebral ischemic conditions to be distinguished from intracranial hemorrhage, likely explaining the clearer risk signal for the former and extended to associated conditions such as diabetes and chronic kidney disease. Importantly and in contrast with all previously cited studies, we addressed potential confounding by indication directly by restricting the comparison to women who experienced infertility prior to their first birth. We complemented the analysis by assessing cardiovascular risks among all women with primary infertility compared to same-age women with no prior record of infertility, and explored the role of differences in baseline.

In 2016 a panel of over 40 international experts called for more population health studies and better use of existing databases to investigate the links between infertility and later cardiometabolic health [26]. Since then, a series of studies from North America have shown women with infertility at elevated risk of premature mortality and morbidity from cardiovascular and metabolic disease [27]. Importantly, similar associations have been observed in men, in which poor semen quality has been linked to increased risk of chronic disease and early mortality [28,29]. These findings underscore that infertility may serve as a broader marker of underlying systemic health risk across sexes, reinforcing the importance of longitudinal follow-up in both women and men undergoing fertility evaluation. Following a recent report from the Trøndelag Health Study in Norway finding moderate associations between infertility and risk of stroke and coronary heart disease [30], the editorial called out limitations of previous studies, including difficulties establishing temporality, insufficient follow-up, and poor infertility surveillance. Nordic register studies have the potential to overcome many of these limitations, yet attempts are few to date. An early follow-up of women giving birth in Sweden between 1983 and 2005 noted an excess risk of cardiovascular disease (coronary heart disease, stroke, and heart failure combined) in women who had given birth after more than 5 years of infertility [31]. Adding almost 20 years of follow-up, our study found that women who experienced infertility before their first birth developed clearly elevated risks of actue myocardial infarction, type 2 diabetes, and heart failure compared to women with no record of primary infertility.

### Strengths and limitations of the study

In summary, our study adds to the existing literature by assessing the long-term cardiovascular risk associated with both ART and infertility which is particularly important for understanding cardiovascular and metabolic outcomes that become more prevalent at older ages. By incorporating multiple comparison groups and applying inverse probability weighting to adjust for baseline differences we were able to isolate the potential contribution of ART from that of the underlying infertility and its associated risk factors. Drawing on a large, population-based cohort with nationwide coverage and ability to follow women 30 years post-conception we provide a robust and clinically relevant evaluation of cardiometabolic risk in this growing patient population.

Key strengths of the study include the use of high-quality national registers with prospectively collected data, enabling us to study a nationwide population with minimal risk of selection bias, recall bias, or reverse causation. Exposure to ART was primarily established from clinical reports, and the experience of infertility was surveyed at enrolment to antenatal care. The careful definition of exposures and outcomes, adjustment for key confounders and the separate evaluation of cerebral ischemic conditions and intracranial hemorrhage represent further strengths. By estimating cumulative incidences starting from conception, we aimed to capture the full risk profile while avoiding potential issues with survival bias or non-proportional hazards.

However, several limitations should be considered. Although we limited the study period to avoidthe earliest years of ART implementation when exposure was very rare and likely also highly selective, the number of women conceiving with ART was still low, and their risks thus estimated with limited precision, reducing the ability to distinguish any observed differences from the null. Careful interpretation is particularly warranted in the beginning and end of follow-up, due to very few events and decreasing numbers at risk respectively. The priority on adequate long-term follow-up further precluded analyses of specific risks such as those associated with frozen embryo transfer, which was only introduced toward the end of the study period. Similar to previous studies we did not have information on failed ART attempts and could only study the exposure in women giving birth. Among women with infertility, there could be some healthy selection of the comparison group (able to conceive without ART), whereby we would expect any observed associations between ART and cardiovascular outcomes to be overestimated. More importantly we do not know if our findings generalize to treatment-resistant women, and further work is required to understand if and how their exposure increases their risk of cardiovascular disease. Prior to 1997 the identification of diabetes was based on ICD-8/9 code 250, which does not distinguish between type 1 and type 2 diabetes. Still, the exclusion of women diagnosed before the start of follow-up should reduce the likelihood of including type 1 diabetes, which typically manifests at younger ages. Diabetes is likely also subject to some misclassification due to inadequate coverage of outpatient care. Expecting such to be non-differential (once the influence of calendar year on ART has been accounted for) and thus bias findings toward the null, we note that a clear difference in risk could still be observed in the comparison of women with and without infertility. We also note that since we did not have access to primary care records, the findings should generally reflect more severe or co-morbid cases. Finally, although constrained by the availability and historical coverage of register data, the inclusion of measured background characteristics and the use of multiple comparison groups were sufficient to explain the observed excess risk in women who conceived with ART.

### Clinical implications

The findings of this study have important clinical implications. First and most important, while largely reassuring to providers and patients the findings also suggest some need for caution regarding the long-term cardiovascular risk of ART use, and a need for further study of the possible long-term risks of ischemic cerebral conditions. Second, the additional assessment of overall risks to women experiencing primary infertility adds to a growing body of literature on the link between infertility and later cardiovascular health, recognized in a scientific statement by the American Heart Association in January 2025 [32]. Our study, featuring the longest follow-up to date, provides strong epidemiological support for this recognition. Of note, the clinical definition of infertility (time to pregnancy > 1 year) that we relied on captures the couple experience of infertility, irrespective of its root causes. Hypothesizing that the observed cardiovascular risks are primarily due to causes and risk factors of infertility, we may expect greater risks associated with female factor infertility. Beyond the scope of our study, further research to improve risk prediction and/or understand mechanisms should consider the roles of specific types of infertility and their risk factors.

When evaluating women’s cardiovascular risk, cardiologists should consider the reproductive history, including potential infertility. Conversely, whenever infertility emerges in the clinical history it should prompt targeted cardiovascular evaluation. Women with infertility may be offered targeted cardiovascular screenings for hypertension, dyslipidemia, and insulin resistance to allow early detection of subclinical risk factors and timely intervention to reduce the long-term cardiovascular morbidity. Typically, infertility is first detected by health care if/when the woman seeks fertility assistance. To integrate this care with cardiological expertise could optimize cardiovascular risk management, by ensuring that underlying infertility-related risk factors, such as endothelial dysfunction, hypertension, or insulin resistance, are properly identified and managed, before, during, and after the fertility assistance.

## CONCLUSIONS

The findings of this study adds to the growing evidence that underlying factors connected to infertility compound long-term risk of cardiovascular events, and while use of ART appears largely safe it may increase the risk of developing ischemic cerebral conditions later in life. The findings underscore the importance of monitoring cardiovascular health in women with infertility, as the underlying causes of infertility in particular may contribute to a higher risk of cardiovascular disease. Further research is warranted to explore the mechanisms linking infertility with cardiovascular outcomes and the specific risk of ischemic cerebral conditions following use of ART, to establish appropriate screening and preventive strategies for these population.

## SUPPORTING INFORMATION LEGENDS

### Methods

S1 Table. Codes used to identify exposure.

S2 Table. Codes used to identify and define outcomes.

S1 Fig. Directed acyclic graph of hypothesized relationships.

S2 Fig. Multiple imputation diagnostics: (A) MCMC convergence trace plots for the means and standard deviations of imputed covariates; (B) density plots of observed and imputed distribution of BMI; and (C) strip plots of observed and imputed values of BMI.

S3 Table. Multiple imputation diagnostics: Observed and imputed distributions of categorical covariates.

S4 Fig. Mirror density plot of pooled propensity scores.

### Results

S5 Table. Cumulative failure proportions at 10, 20, and 30 years, risk differences, and risk ratios for the fully-weighted comparison of ART and no ART among women with primary infertility.

S5 Fig. Kaplan–Meier failure curves of chronic kidney disease and aortic aneurysm or dissection for the fully-weighted comparison of ART and no ART among women with primary infertility.

S6 Table. Cumulative failure proportions at 10, 20, and 30 years, risk differences, and risk ratios for cerebral ischemic conditions.

S6 Fig. Kaplan–Meier failure curves of the main cardiovascular outcomes for age-weighted comparison of women with and without primary infertility.

S7 Table. Cumulative failure proportions at 10, 20, and 30 years, risk differences, and risk ratios for the fully-weighted comparison of women with and without primary infertility.

S7 Fig. Kaplan–Meier failure curves of chronic kidney disease and aortic aneurysm or dissection for fully-weighted comparison of women with and without primary infertility.

S8 Fig. Kaplan–Meier failure curves of cardiovascular diseases for fully-weighted comparison of ICSI and no ART among women with primary infertility.

## FIGURE TITLES AND LEGENDS

### Figure legends

**S1 Fig**

ART: Assisted reproductive technology; CVD: Cardiovascular disease; BMI: Body mass index; Origin: Country or region of origin; Genes: Genetic factors; Diseases: Pre-existing chronic diseases/comorbidities; Infertility: Infertility status; SEI: Socioeconomic Index; Age: Maternal age.

**S3 Table**

Continuous variables were modeled with cubic splines.

## REFERENCES

1. Jain M, Singh M. Assisted Reproductive Technology (ART) Techniques. StatPearls. Treasure Island (FL): StatPearls Publishing; 2025. Available: http://www.ncbi.nlm.nih.gov/books/NBK576409/

2. Baker VL, Dyer S, Chambers GM, Keller E, Banker M, de Mouzon J, et al. International Committee for Monitoring Assisted Reproductive Technologies (ICMART): world report for cycles conducted in 2017-2018. Hum Reprod. 2025;40: 1110–1126. doi:10.1093/humrep/deaf049

3. Udell JA, Lu H, Redelmeier DA. Long-term cardiovascular risk in women prescribed fertility therapy. J Am Coll Cardiol. 2013;62: 1704–1712. doi:10.1016/j.jacc.2013.05.085

4. Crandall CJ, Barrett-Connor E. Endogenous sex steroid levels and cardiovascular disease in relation to the menopause: a systematic review. Endocrinol Metab Clin North Am. 2013;42: 227–253. doi:10.1016/j.ecl.2013.02.003

5. Zhang J, Xu J-H, Qu Q-Q, Zhong G-Q. Risk of Cardiovascular and Cerebrovascular Events in Polycystic Ovarian Syndrome Women: A Meta-Analysis of Cohort Studies. Front Cardiovasc Med. 2020;7: 552421. doi:10.3389/fcvm.2020.552421

6. Dayan N, Joseph KS, Fell DB, Laskin CA, Basso O, Park AL, et al. Infertility treatment and risk of severe maternal morbidity: a propensity score-matched cohort study. CMAJ. 2019;191: E118–E127. doi:10.1503/cmaj.181124

7. Henriksson P, Westerlund E, Wallén H, Brandt L, Hovatta O, Ekbom A. Incidence of pulmonary and venous thromboembolism in pregnancies after in vitro fertilisation: cross sectional study. BMJ. 2013;346: e8632. doi:10.1136/bmj.e8632

8. Olausson N, Discacciati A, Nyman AI, Lundberg F, Hovatta O, Westerlund E, et al. Incidence of pulmonary and venous thromboembolism in pregnancies after in vitro fertilization with fresh respectively frozen-thawed embryo transfer: Nationwide cohort study. J Thromb Haemost. 2020;18: 1965–1973. doi:10.1111/jth.14840

9. Westerlund E, Brandt L, Hovatta O, Wallén H, Ekbom A, Henriksson P. Incidence of hypertension, stroke, coronary heart disease, and diabetes in women who have delivered after in vitro fertilization: a population-based cohort study from Sweden. Fertil Steril. 2014;102: 1096–1102. doi:10.1016/j.fertnstert.2014.06.024

10. Dayan N, Filion KB, Okano M, Kilmartin C, Reinblatt S, Landry T, et al. Cardiovascular Risk Following Fertility Therapy: Systematic Review and Meta-Analysis. J Am Coll Cardiol. 2017;70: 1203–1213. doi:10.1016/j.jacc.2017.07.753

11. Strain T, Wijndaele K, Sharp SJ, Dempsey PC, Wareham N, Brage S. Impact of follow-up time and analytical approaches to account for reverse causality on the association between physical activity and health outcomes in UK Biobank. Int J Epidemiol. 2020;49: 162–172. doi:10.1093/ije/dyz212

12. Sattar N, Preiss D. Reverse Causality in Cardiovascular Epidemiological Research: More Common Than Imagined? Circulation. 2017;135: 2369–2372. doi:10.1161/CIRCULATIONAHA.117.028307

13. Simons PIHG, Cornelissen MEB, Valkenburg O, Onland-Moret NC, van der Schouw YT, Stehouwer CDA, et al. Causal relationship between polycystic ovary syndrome and coronary artery disease: A Mendelian randomisation study. Clin Endocrinol (Oxf). 2022;96: 599–604. doi:10.1111/cen.14593

14. Udell JA, Lu H, Redelmeier DA. Failure of fertility therapy and subsequent adverse cardiovascular events. CMAJ. 2017;189: E391–E397. doi:10.1503/cmaj.160744

15. Ludvigsson JF, Otterblad-Olausson P, Pettersson BU, Ekbom A. The Swedish personal identity number: possibilities and pitfalls in healthcare and medical research. Eur J Epidemiol. 2009;24: 659–667. doi:10.1007/s10654-009-9350-y

16. Cnattingius S, Källén K, Sandström A, Rydberg H, Månsson H, Stephansson O, et al. The Swedish medical birth register during five decades: documentation of the content and quality of the register. Eur J Epidemiol. 2023;38: 109–120. doi:10.1007/s10654-022-00947-5

17. Wettergren B, Blennow M, Hjern A, Söder O, Ludvigsson JF. Child Health Systems in Sweden. J Pediatr. 2016;177S: S187–S202. doi:10.1016/j.jpeds.2016.04.055

18. Ludvigsson JF, Andersson E, Ekbom A, Feychting M, Kim J-L, Reuterwall C, et al. External review and validation of the Swedish national inpatient register. BMC Public Health. 2011;11: 450. doi:10.1186/1471-2458-11-450

19. Brooke HL, Talbäck M, Hörnblad J, Johansson LA, Ludvigsson JF, Druid H, et al. The Swedish cause of death register. Eur J Epidemiol. 2017;32: 765–773. doi:10.1007/s10654-017-0316-1

20. Eriksson A, Stenlund H, Ahlm K, Boman K, Bygren LO, Johansson LA, et al. Accuracy of death certificates of cardiovascular disease in a community intervention in Sweden. Scand J Public Health. 2013;41: 883–889. doi:10.1177/1403494813499653

21. Austin PC, White IR, Lee DS, van Buuren S. Missing Data in Clinical Research: A Tutorial on Multiple Imputation. Can J Cardiol. 2021;37: 1322–1331. doi:10.1016/j.cjca.2020.11.010

22. Toutenburg H. Rubin, D.B.: Multiple imputation for nonresponse in surveys/. Statistical Papers. 1990;31: 180–180. doi:10.1007/BF02924688

23. von Elm E, Altman DG, Egger M, Pocock SJ, Gøtzsche PC, Vandenbroucke JP, et al. The Strengthening the Reporting of Observational Studies in Epidemiology (STROBE) statement: guidelines for reporting observational studies. J Clin Epidemiol. 2008;61: 344–349. doi:10.1016/j.jclinepi.2007.11.008

24. Magnus MC, Fraser A, Håberg SE, Rönö K, Romundstad LB, Bergh C, et al. Maternal Risk of Cardiovascular Disease After Use of Assisted Reproductive Technologies. JAMA Cardiol. 2023;8: 837–845. doi:10.1001/jamacardio.2023.2324

25. Pivato CA, Inversetti A, Condorelli G, Chieffo A, Levi-Setti P emanuele, Latini A chiara, et al. Cardiovascular safety of assisted reproductive technology: a meta-analysis. European Heart Journal. 2024. doi:10.1093/eurheartj/ehae886

26. Cedars MI, Taymans SE, DePaolo LV, Warner L, Moss SB, Eisenberg ML. The sixth vital sign: what reproduction tells us about overall health. Proceedings from a NICHD/CDC workshop. Hum Reprod Open. 2017;2017: hox008. doi:10.1093/hropen/hox008

27. Murugappan G, Li S, Lathi RB, Baker VL, Eisenberg ML. Increased risk of incident chronic medical conditions in infertile women: analysis of US claims data. Am J Obstet Gynecol. 2019;220: 473.e1–473.e14. doi:10.1016/j.ajog.2019.01.214

28. Latif T, Kold Jensen T, Mehlsen J, Holmboe SA, Brinth L, Pors K, et al. Semen Quality as a Predictor of Subsequent Morbidity: A Danish Cohort Study of 4,712 Men With Long-Term Follow-up. Am J Epidemiol. 2017;186: 910–917. doi:10.1093/aje/kwx067

29. Eisenberg ML, Li S, Behr B, Cullen MR, Galusha D, Lamb DJ, et al. Semen quality, infertility and mortality in the USA. Hum Reprod. 2014;29: 1567–1574. doi:10.1093/humrep/deu106

30. Skåra KH, Åsvold BO, Hernáez Á, Fraser A, Rich-Edwards JW, Farland LV, et al. Risk of cardiovascular disease in women and men with subfertility: the Trøndelag Health Study. Fertil Steril. 2022;118: 537–547. doi:10.1016/j.fertnstert.2022.05.038

31. Parikh NI, Cnattingius S, Mittleman MA, Ludvigsson JF, Ingelsson E. Subfertility and risk of later life maternal cardiovascular disease. Hum Reprod. 2012;27: 568–575. doi:10.1093/humrep/der400

32. Mauricio R, Sharma G, Lewey J, Tompkins R, Plowden T, Rexrode K, et al. Assessing and Addressing Cardiovascular and Obstetric Risks in Patients Undergoing Assisted Reproductive Technology: A Scientific Statement From the American Heart Association. Circulation. 2025;151: e661–e676. doi:10.1161/CIR.0000000000001292

